# Re-examining the meningitis belt: associations between environmental factors and epidemic meningitis risk across Africa

**DOI:** 10.1101/2025.02.25.25322867

**Authors:** Molly Cliff, Sally Jahn, Andre Bita Fouda, Anderson Latt, Clement Lingani, Caroline Trotter

## Abstract

In 2002 Molesworth et al created a logistic regression model to investigate the spatial distribution of meningitis epidemics in Africa and determine geographical risk. We aim to understand if the meningitis belt geography and environmental risk factors have changed in the last 20 years by updating this analysis. Epidemic bacterial meningitis data from 2003-2022 were provided by WHO-AFRO. Districts (ADMN2) across Africa were coded as 1 if they had experienced a meningitis outbreak and 0 if they hadn’t. Environmental data were obtained from open sources. To preserve seasonal climate variation, monthly means of wind speed, rainfall, dust, and humidity were processed into climatic profiles using k-means clustering. Logistic regression was carried out with meningitis epidemic history as the dependent variable and k-means clusters of rainfall, dust, humidity, and windspeed, alongside average land-cover type as the independent variables. A sensitivity analysis was conducted excluding the Democratic Republic of Congo (DRC), due to limited laboratory confirmation of suspected cases. Rainfall, dust, and humidity demonstrated the strongest statistical association with meningitis outbreaks and were included in our final model. With a probability cutoff value of >0.4, our model had a specificity and sensitivity of 81.0% and 84.3% respectively in identifying districts having experienced a meningitis epidemic. With an extended dry season, the Sahelian region had the highest risk of meningitis outbreaks (probability >0.8). The inclusion/exclusion of the DRC had a significant impact on our model. Whilst Republic of the Congo, Gabon, Liberia, and Angola had a moderate risk of meningitis (probability >0.4) within our full model, no countries surrounding the meningitis belt were at risk for outbreaks within the sensitivity analysis. Although our research may suggest a south-westerly expansion of the meningitis belt this is difficult to verify. Surveillance of suspected meningitis cases should be accompanied by laboratory confirmation to improve outbreak definition.

## Introduction

The African meningitis belt, running from Ethiopia to Senegal, has a high seasonal incidence of bacterial meningitis comprising approximately 80% of all meningitis cases within Africa [1].The meningitis belt was first defined in 1963 by Lapeyssonie, as an area within sub-Saharan Africa including 11 countries with a high incidence of meningitis epidemics with approximately 300-1100 mm isohyets of annual rainfall [2]. Lapeyssonie additionally noted that meningitis outbreaks increasingly occurred during the warmer, dry season within sub-Saharan Africa, highlighting a correlation with seasonal weather patterns [2]. In 1971 an extension of the meningitis belt towards East Africa was proposed to include regions of Rwanda, Burundi, Uganda, Tanzania and Kenya based on observation of epidemics from 1911-1965 [3]. In 1981 Greenwood furthered this work to include areas of 17 countries, with western and eastern extensions from Lapeyssonie and the exclusion of Democratic Republic of Congo (DRC) [4]. Currently, with further input from Molesworth et al and risk assessments performed in the context of the Meningitis Vaccine Project, the meningitis belt spans 26 countries. It is estimated that nearly 80% of all meningitis cases within Africa occur within the belt, with the region comprising over 45% of cases globally in 2016 [5].

Seasonality and climate variability have a key influence on the spatiotemporal distribution of bacterial meningitis. Within the December to April dry season, initially dominated by the Harmattan winds and increases in dust concentration, disease incidence can reach 800 cases per 100,000 people [6]. Bacterial meningitis incidence decreases with the onset of the wet season characterized by pre-monsoon rainfall [7]. The co-occurrence of meningitis cases – predominantly due to *Neisseria meningitidis* - and climate variability within the belt suggests a relationship between meningitis incidence and climate [1] Several climatic models have been able to highlight the spatiotemporal relationship between meningitis cases, and meteorological weather events[8]. Chen et al established a connection between increased temperature variability, and heightened global meningitis risk [9], while Tall et al found a significant association between meningitis outbreaks and both relative humidity and temperature when forecasting meningitis cases in Burkina Faso[7]. Several additional studies within the meningitis belt have concluded that humidity, dust, and rainfall are associated with meningitis epidemics [10,11,12,13]. The specific mechanism is not known, but seasonal climatic factors may aid in the promotion of bacterial transmission and progression from carriage to invasive disease [14,15]. Work linking environmental factors and meningitis has been incorporated in early warning systems and prevention and has been used successfully to support awareness, preparedness and response actions for meningitis outbreaks [12,13,16]

A 2002 study by Molesworth et al investigated the spatial distribution of meningitis epidemics in Africa occurring between 1980 and 1999 to define the geographical risk of meningitis across Africa. This determined that in addition to those countries within the meningitis belt at the time Guinea-Bissau, Guinea, Côte d’Ivoire, Togo, the Central African Republic and Eritrea had a relatively high epidemic burden [17]. This work was then expanded into a model examining the relationship between epidemic location and environmental variables to locate districts within Africa with increased risk of meningitis epidemics. Molesworth et al determined that absolute humidity and land-cover type were able to best predict meningitis epidemics within their logistic regression model. Additionally, dust and rainfall zonal profiles were also independently associated with epidemic location [18]. The association with dust is thought to be in line with the increase in dust particles following the Sahelian droughts of the 1970s and 1980s [19]. The model also demonstrated that climatic zones without distinctive seasonal changes including desert and forest areas are less likely to have epidemics as contrasted to regions with wet and dry seasons. [18]. Molesworth et al’s work was informative for health policy and surveillance activities and drove monitoring of climate and environmental change on epidemic risk across Africa. In 2006, Savory et al sought to evaluate Molesworth et al’s risk model, examining 71 meningitis outbreaks from 2000-2004, suggesting that epidemics may be extending southwards within the Sahelian region. The proposed expansion of the meningitis belt in line with this, towards Togo, the Central African Republic, Cameroon, and Côte d’Ivoire was consistent with regional environmental changes [20]. Alterations in land usage, combined with climatic factors, potentially led to a humidity reduction combined with an increase in dust, favouring epidemic conditions.

Our work will present an update on the associations between environmental variables and meningitis epidemic risk for meningitis across Africa. Despite the changing epidemiology of bacterial meningitis since 2000 due to the introduction of Hib, pneumococcal and particularly meningococcal vaccines, environmental risk factors for outbreaks remain consistent and relevant within the context of widespread vaccination. The introduction of MenAfriVac from 2010 onwards in mass immunisation campaigns reduced the meningitis burden in vaccinated countries, and whilst outbreaks of group A meningococcal (NmA) epidemics have decreased as a result [21] there have been persistent outbreaks due to other meningococcal groups (NmW, NmX and NmC) as well as pneumococcal meningitis since 2013 [22].

Understanding the evolving impact of climate change on meningitis incidence is vital, given that more than half of known human pathogenic diseases can be exacerbated by climate change [23]. Now that 20 years have passed since the original publication, we aim to understand if the geography of the meningitis belt and risk factors associated with meningitis have changed. Due to increasing data availability, we can analyse over 20 years of meteorological data, allowing for a focus on climate dynamics, as opposed to shorter-term weather assessments. This enables the isolation and identification of climatic trends amidst the natural variability of weather. Additionally, with both the quality and resolution of environmental data having significantly improved since 2002, updating previous work should serve to create a more reliable risk model. By repeating the methods used by Molesworth et al, with new data to look for a signal of change before going on to investigate the relationship between climate and meningitis further, we can ensure that any observed changes are due to the data used rather than the methodology. This analysis is highly relevant to understanding of the effect of climate on meningitis incidence in the context of a changing climate.

## Methods

### Epidemiological data

We included meningitis epidemics reported to the WHO enhanced meningitis surveillance system [24] from 2003 through to 2022. The enhanced meningitis surveillance system reports disease data from 24 countries across sub-Saharan Africa which are either in or adjacent to the meningitis belt and are at high risk of meningitis outbreaks. Whilst this excludes countries in North Africa, that have been previously identified as high-risk, outbreaks within this region during our study period have been limited [25]. We defined an epidemic as any district reaching the epidemic threshold of 10 suspected meningitis cases per 100,000 population per week or with a seasonal cumulative incidence of 100 suspected meningitis cases per 100,000 population [26]. Outbreak metadata included the year and ADMN2 district of each meningitis epidemic as well as population size and attack rate. As pathogen data on individual outbreaks was unavailable we did not examine the impact of vaccination as part of our main investigation. Many outbreaks would have occurred before meningococcal and pneumococcal vaccinations were introduced and the risk of epidemics from other serogroups/ serotypes remains. Meningitis epidemics were assigned to their nearest ADMN2 district within a GADM shapefile using Stata/ SE 18 [27]. Due to the changing nature of district boundaries, epidemics reported to older administrative boundaries were mapped to existing (2024) ADMN2 districts using a WHO Polio shapefile in R 4.3.1. We calculated ADMN2 level district intersections between the WHO Polio and GADM shapefiles. Intersections between the two shapefiles were filtered where they overlap by 50% and included as an outbreak district. We then repeated this for shapefiles overlapping by 30% to increase the number of outbreaks assigned to a GADM ADMN2 district.

### Environmental data

Publicly available environmental data with uniform latitude-longitude grid-based coverage was used in this analysis. Variables used were originally derived from the interpolation of climatic anomalies across global weather station networks (Table 1).

**Table 1.**
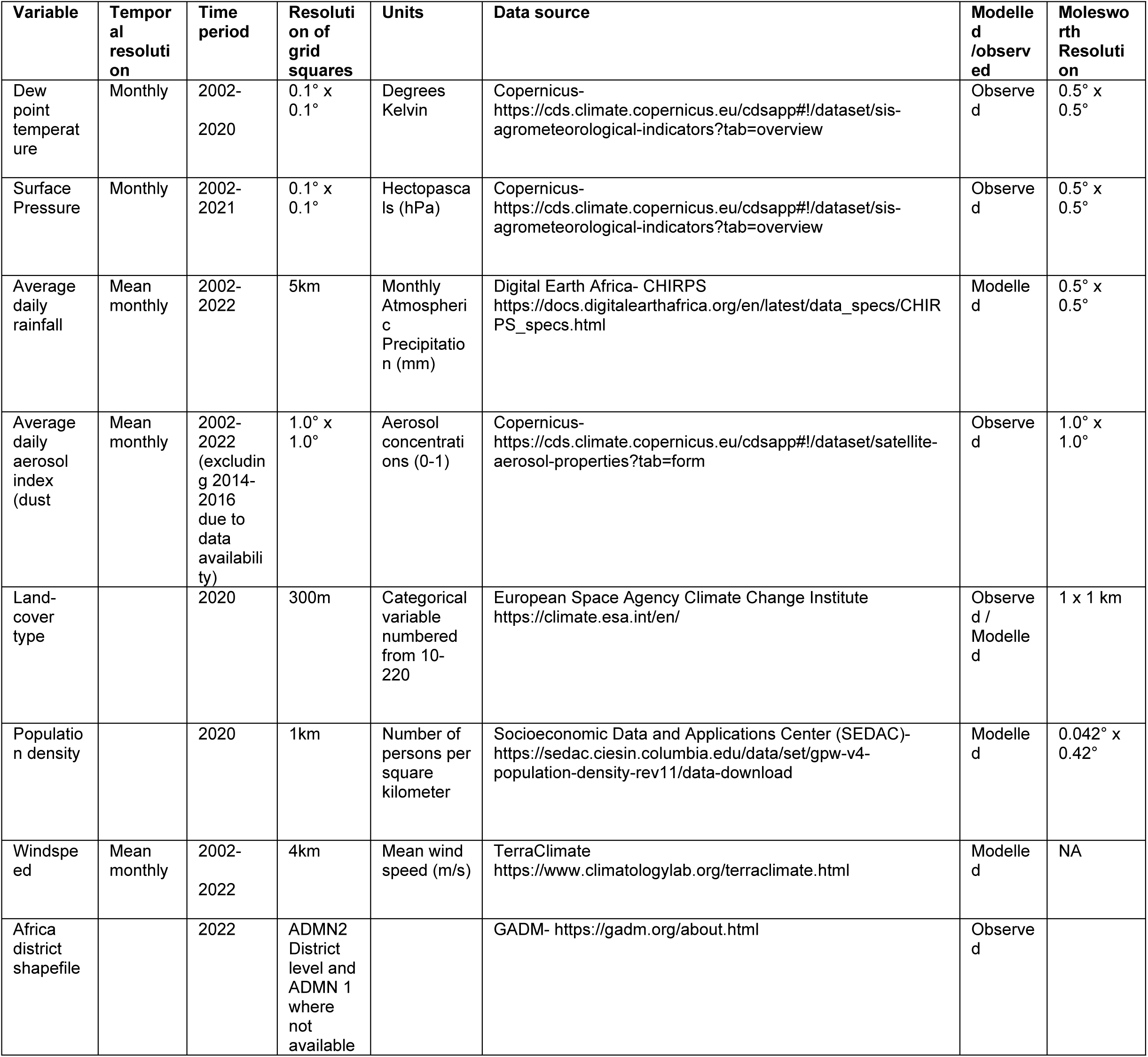
Characteristics of environmental variables to be included within risk modelling as compared with Molesworth et al.

We examined the impact of specific humidity (g/m3), average daily aerosol index (dust) (aerosol concentrations 0-1), rainfall (monthly atmospheric precipitation-mm), population density (number of people per square kilometer) and land coverage (categorical land cover class) on meningitis epidemics. By isolating environmental determinates of outbreaks we aim to better understand broader climate and environment patterns influencing the risk of meningitis. Specific humidity was derived from dew point temperature and surface pressure using the following formula [28].

1. Calculating saturation water vapor pressure (*es*) from Teten’s formula:

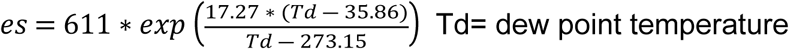

2. Calculating specific humidity

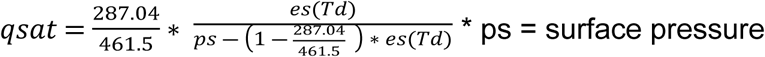

Whilst Molesworth et al used absolute humidity within their analysis, we opted for specific humidity so that we could apply our model to future variable Global Circulation Models as part of a future risk mapping project. We also examined the effect of wind speed on meningitis epidemics, a variable not originally studied in Molesworth’s work. When determining which environmental data sources to use in the model we prioritized data resolution. We consistently used gridded datasets in their native spatial resolution and subsequently calculated area-level estimates for each GADM unit based on these inputs. Due to the high resolution of rainfall, landcover and windspeed data we resampled raster data in R 4.3.1, using bilinear interpolation to effectively decrease raster resolution without compromising data quality. Environmental variables and their respective data sources used can be seen in Table 1.

To preserve seasonal climate variation, while reducing data dimensionality, the monthly means of each temporal variable were processed into a single surface comprising of climatic variable profiles, as in Molesworth’s original methodology. We decreased the data dimensionality of temporal variables due to the size of the raster data set which would necessitate a high level of computing power. Principle component analysis was carried out within R to decrease both the data dimensions and the number of raster layers to be analysed, whilst retaining 95% of the seasonal information retained within the raster data. The K means algorithm was then used to divide each seasonal variable raster into distinct clusters based on data similarity. This is achieved through the K-means algorithm’s ability to assign data points to clusters whilst continuously updating the clusters’ respective mean values, allowing the algorithm to minimize the intra-cluster variance. This follows the same methodology implemented by the ADDAPIX software within Molesworth et al’s original analysis. We used both model iteration and the elbow method to define the optimal number of clusters for each environmental variable to use in a k-means clustering algorithm [29,30,31]. Following on from this we carried out an initial visual check against Koppen Geiger and Schultz climate zonal maps alongside Molesworth et al’s environmental clustering to determine the appropriateness and success of environmental variable clustering [32].

Finally, for each of the 6350 districts within Africa, the extract function created an average value for each environmental variable within the GADM continental shapefile based on the native resolution of each dataset apart from where previously resampled. ADMN2 districts were used for environmental analysis in all countries where available, apart from in Libya, Western Sahara and Comoros where only ADMN1 level subdivisions are available via GADM. Any environmental clusters with less than 50 districts were assigned to their nearest neighbour environmental cluster using a cluster dendrogram to prevent complete separation and inflated significance. Figure 1 demonstrates the results of our environmental clustering profiles for rainfall, aerosol optical depth(AOD) and specific humidity. For consistency with Molesworth et al’s analysis, we used logistic regression to allow for direct comparison between risk maps. By using logistic regression we can examine the impact of environmental risk factors across the whole of the meningitis belt and its surrounding area.

**Figure 1:** Seasonal patterns within the different cluster profiles of aerosol optical depth (AOD) (1a), rainfall (1b) and specific humidity (1c) (right panel) alongside the relative assigned district classes (left panel).

Within our study, the dependent variable was ADMN district history of meningitis epidemic, whilst the independent variables were k-means clusters of rainfall, aerosol optical depth, specific humidity and windspeed, and average landcover type and population density. Due to our use of k-means clustering as opposed to using raw variables, there were no confounding variables present. We ran the model weighting for inverse district size, population density and growth and fertility rate [33]. We weighted against larger districts as these are less likely to be appropriately represented by one cluster class due to ecological variation. In weighting for population growth and fertility rate we aimed to capture the relationship between meningitis outbreaks and population size through accounting for population change over time. When accounting for population weighting we initially used a pixel-based weighting system to derive average population density for each of the 6350 ADMN2 districts across Africa. This was achieved through using SEDAC population estimates, given every 5 years from 2000-2020 [34] to derive population density for each ADMN2 district over the study time frame. Additionally, we obtained country-level population density, fertility rate, and population growth from UNPOP’s 2022 dataset [33]. We manually balanced the class weights for districts with/without epidemics by prioritising the f1 score, improving model precision and recall. This was carried out due to the large imbalance between ADMN2 districts with/without an epidemic. Area averages were also based on a cosine of latitude weighting due to our use of regular longitude/latitude gridded data. We also conducted a sensitivity analysis, running the logistic regression model without the Democratic Republic of Congo, due to the country’s very limited laboratory confirmation of suspected cases [25].

### Risk Analysis

Backwards stepwise logistical regression was used to identify the risk of meningitis outbreak within a district in Africa and the relative environmental associations across the whole dataset. For environmental variables, we used a threshold significance level of <0.05 when identifying statistically significant associations. The probability of each district ever having had an epidemic was predicted through the model, before being grouped into risk categories and then mapped. The model was tested by checking specificity and sensitivity, examining collinearity through the Variance inflation factor, as well as utilising McFadden’s Pseudo-R2 Interpretation for goodness of fit. Finally, a receiver-operator characteristics (ROC) curve was used to examine the model’s capability to discriminate between true positives and false positives across all possible thresholds. The logistic regression model’s ability to predict outbreaks was verified using mean validation, splitting the data into “train” (80% of the data) and “test” (20% of the data) sections. Models were recreated with the train data 10 times and then used to predict epidemic risk within the test data. Mean sensitivity and specificity of the validation data were compared against that of the backward stepwise model formed from the whole data set. R code used to run this analysis can be found within the following GitHub repository (https://github.com/molly-cliff/Meningitis-belt-location).

## Results

Between 2002 and 2022 there were 1069 reported district level epidemics with 10 suspected cases per 100,000 population per week alongside 14 district-level annual outbreaks with at least a seasonal cumulative incidence of 100 suspected meningitis cases per 100,000 population. 55 epidemics were dropped from the final dataset as we were unable to assign them to an appropriate ADMN2 district in either the WHO Polio or GADM shapefile. After considering duplicate districts (with more than one epidemic) there were 523 ‘ever affected’ districts representing 8.2% of all 6350 districts in Africa (Fig 2).

**Figure 2:** Map of Africa showing location of meningitis epidemics between 2003-2022.

Epidemics most prominently occurred within the Central Sahelian region of Africa, with Burkina Faso, Chad and Niger having over 50% of their ADMN2 districts affected by an outbreak. Additionally, within this region, Nigeria had over 200 districts affected by a meningitis outbreak. However, across the most western and eastern parts of the Sahel, Mauritania and Sudan both reported zero outbreaks with Mali also reporting a low number of affected districts. Across the remainder of the meningitis belt Togo, Ghana, The Gambia, and Ethiopia were also affected by meningitis outbreaks. Whilst the north of the Democratic Republic of Congo was thought to be the region of the country within the meningitis belt, between 2002 and 2022 outbreaks occurred throughout the country. Within our study period, 51% of all districts within the DRC experienced a meningitis outbreak. If districts in DRC are excluded then there were 442 ever-affected epidemic districts in our dataset.

Specific humidity, rainfall, windspeed, aerosol optical depth and landcover category were independently associated with the location of epidemics. However, we found that specific humidity, rainfall, and aerosol optical depth profiles were the best predictors within our multivariate model. Backwards stepwise logistic regression suggested the removal of windspeed and landcover as predictor variables due to the comparatively lower association with epidemic location, leading to them being discarded from the final model. Rainfall, aerosol optical depth and humidity were used as independent variables within the final model. Rainfall, and humidity were particularly important as these cluster profiles were able to identify seasonal patterns strongly associated with the meningitis belt, which were statistically associated with outbreak incidence. Weighting for district size, population density, and fertility rate did not improve the model performance and was therefore discarded. In manually balancing the class weighting for the response variable through optimising the f1 score, a weighting of 0.8725 for districts with an epidemic against a weighting of 0.1275 for districts without an epidemic was most appropriate.

Our final backwards stepwise model including the DRC is outlined in Table 2 in relation to its estimated coefficients, standard errors, and variable contribution. Reference clusters were chosen in order to balance standard error across the logistic regression model. We aimed to avoid the risk of inflated standard errors which would undermine model precision, instead choosing clusters based on the dependent variable distribution. The most important factor associated with the distribution of epidemics was rainfall. Collinearity between different variables was largely limited due to the use of k-means clustering analysis instead of directly comparing climatic variables against each other (Fig 3).

**Figure 3:** A matrix demonstrating Pearson’s correlation coefficient representing the normalised covariance between climatic variables used within logistic regression (−1 to 1)

**Table 2:** Logistic regression results regarding association between meningitis outbreaks and rainfall, aerosol optical depth and specific humidity.

| Variable | Epidemic experience<br>(n districts) |  | Multi-variable analysis |  |  |
| --- | --- | --- | --- | --- | --- |
|  | Ever (n) | Never (n) | Odds ratio | 95%<br>Confidence<br>Interval | P value |
| Aerosol optical<br>depth profile |  |  |  |  |  |
| Class 1 | 42 | 388 | 1.38 | 0.66 -2.93 | 0.39 |
| Class 2 | 76 | 1215 | Reference |  |  |
| Class 3 | 20 | 1448 | 0.15 | 0.06-0.40 | < 0.001 |
| Class 4 | 19 | 938 | 0.30 | 0.13 -0.69 | 0.005 |
| Class 5 | 186 | 598 | 0.67 | 0.37-1.21 | 0.18 |
| Class 6 | 8 | 199 | 0.82 | 0.24-2.77 | 0.74 |
| Class 7 | 165 | 475 | 0.99 | 0.49-2.01 | 0.99 |
| Class 8 | 7 | 466 | 0.19 | 0.07-0.51 | < 0.001 |
| Specific<br>humidity profile |  |  |  |  |  |
| Class 1 | 0 | 187 | 0.31 | 0.01-6.84 | 0.46 |
| Class 2 | 5 | 837 | 0.29 | 0.09-0.98 | 0.05 |
| Class 3 | 3 | 143 | 0.86 | 0.17-4.26 | 0.85 |
| Class 4 | 82 | 1181 | 0.96 | 0.49-1.88 | 0.91 |
| Class 5 | 15 | 862 | 0.90 | 0.24-3.43 | 0.87 |
| Class 6 | 19 | 568 | 1.09 | 0.46-2.62 | 0.84 |
| Class 7 | 66 | 102 | 2.17 | 0.82-5.72 | 0.11 |
| Class 8 | 8 | 1267 | 0.15 | 0.05-0.41 | < 0.001 |
| Class 9 | 61 | 376 | Reference |  |  |
| Class 10 | 264 | 304 | 2.35 | 1.21-4.57 | 0.01 |
| Rainfall profile |  |  |  |  |  |
| Class 1 | 365 | 484 | 5.11 | 2.58- 10.13 | < 0.001 |
| Class 2 | 77 | 1057 | Reference |  |  |
| Class 3 | 19 | 1174 | 0.46 | 0.17-1.26 | 0.12 |
| Class 4 | 11 | 1762 | 0.28 | 0.12-0.65 | 0.003 |
| Class 5 | 12 | 636 | 1.27 | 0.32-5.03 | 0.73 |
| Class 6 | 39 | 714 | 0.87 | 0.44-1.72 | 0.70 |

In reference to our risk map and table (Fig 4, Table 3), the central and western part of the Sahelian region of Africa, with lower rainfall, specific humidity and an extended dry season had the highest risk of meningitis outbreaks (probability >0.8), mirroring Molesworth et al’s observations. Burkina Faso, Chad, Gambia, Niger, Nigeria, Senegal, South Sudan and Sudan all had at least 25% of their districts with a very high risk of meningitis outbreaks. Countries to the south of the Sahel, with a shorter and less extreme dry season had a moderate risk of meningitis outbreaks (probability 0.4-0.59). In contrast to Molesworth et al, several countries to the south were identified to have a moderate risk of meningitis outbreaks. Republic of the Congo, Gabon, Liberia, Sierra Leone and Angola all had at least 15% of their districts with a moderate risk of meningitis outbreaks. Notably Gabon and the Republic of Congo had 75.0% and 48.7% districts with a moderate risk of outbreak. These are all countries which border the current meningitis belt. Whilst this may suggest a south-westerly expansion of the meningitis belt more work should be undertaken to verify the reported meningitis outbreaks, specifically with respect to laboratory confirmation of *Neisseria meningitidis*.

**Figure 4:** Risk map of meningitis outbreaks across Africa, based on our final logistical regression results. The key represents the probability of ADMN2 district having experienced a meningitis outbreak based environmental cluster data from 2003-2022.

**Table 3:**
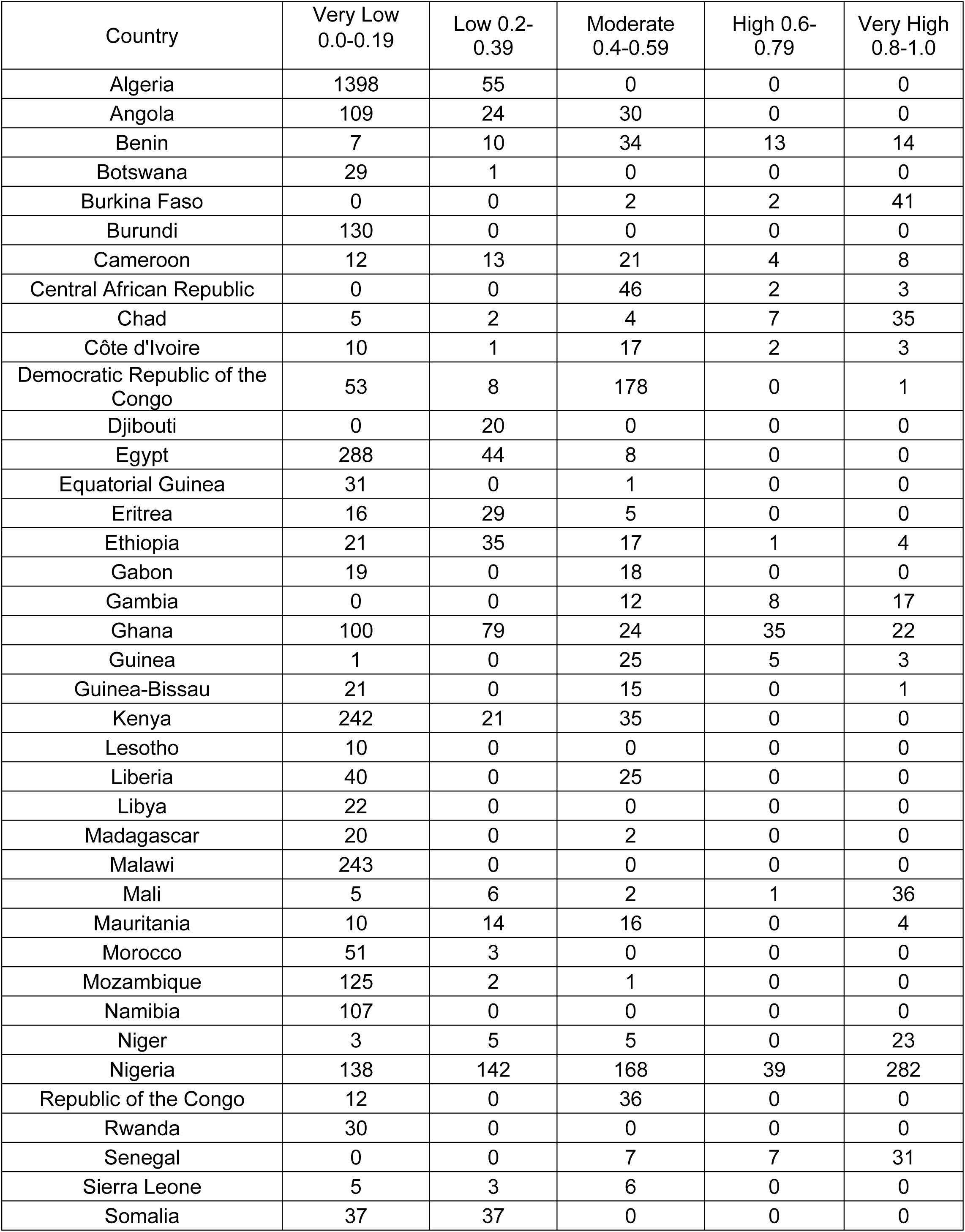
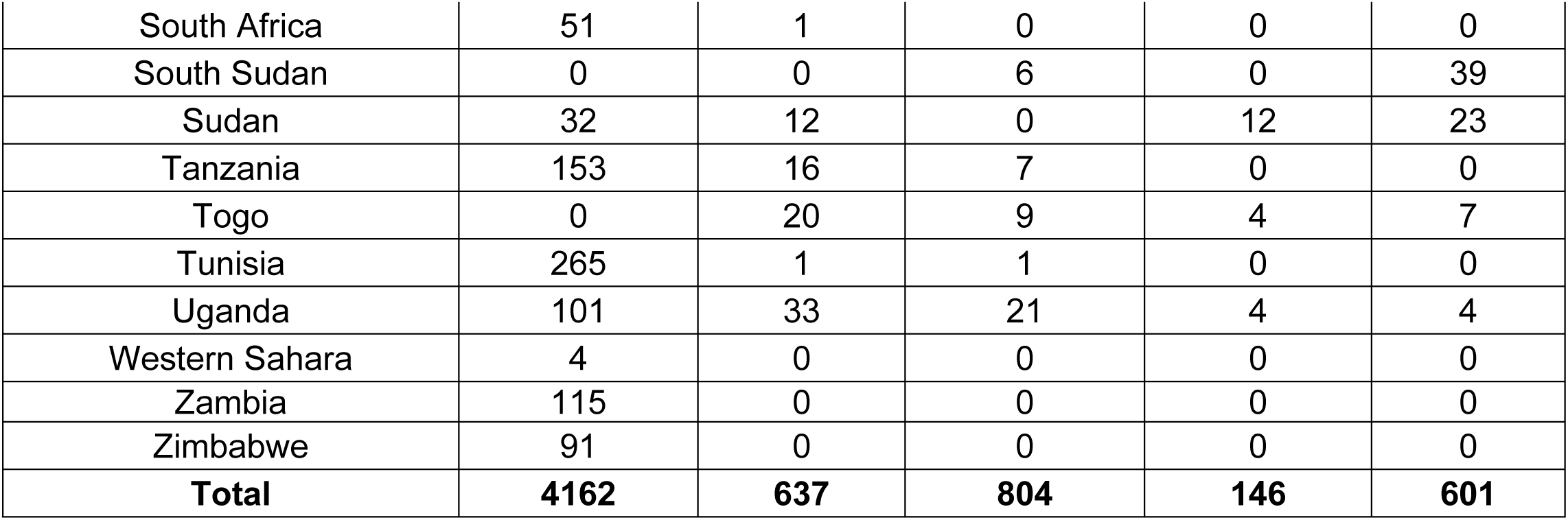
Number of districts within each risk category for all countries across Africa.

| Country | Very Low<br>0.0-0.19 | Low 0.2-0.39 | Moderate<br>0.4-0.59 | High 0.6-0.79 | Very High<br>0.8-1.0 |
| --- | --- | --- | --- | --- | --- |
| Algeria | 1398 | 55 | 0 | 0 | 0 |
| Angola | 109 | 24 | 30 | 0 | 0 |
| Benin | 7 | 10 | 34 | 13 | 14 |
| Botswana | 29 | 1 | 0 | 0 | 0 |
| Burkina Faso | 0 | 0 | 2 | 2 | 41 |
| Burundi | 130 | 0 | 0 | 0 | 0 |
| Cameroon | 12 | 13 | 21 | 4 | 8 |
| Central African Republic | 0 | 0 | 46 | 2 | 3 |
| Chad | 5 | 2 | 4 | 7 | 35 |
| Côte d'Ivoire | 10 | 1 | 17 | 2 | 3 |
| Democratic Republic of the<br>Congo | 53 | 8 | 178 | 0 | 1 |
| Djibouti | 0 | 20 | 0 | 0 | 0 |
| Egypt | 288 | 44 | 8 | 0 | 0 |
| Equatorial Guinea | 31 | 0 | 1 | 0 | 0 |
| Eritrea | 16 | 29 | 5 | 0 | 0 |
| Ethiopia | 21 | 35 | 17 | 1 | 4 |
| Gabon | 19 | 0 | 18 | 0 | 0 |
| Gambia | 0 | 0 | 12 | 8 | 17 |
| Ghana | 100 | 79 | 24 | 35 | 22 |
| Guinea | 1 | 0 | 25 | 5 | 3 |
| Guinea-Bissau | 21 | 0 | 15 | 0 | 1 |
| Kenya | 242 | 21 | 35 | 0 | 0 |
| Lesotho | 10 | 0 | 0 | 0 | 0 |
| Liberia | 40 | 0 | 25 | 0 | 0 |
| Libya | 22 | 0 | 0 | 0 | 0 |
| Madagascar | 20 | 0 | 2 | 0 | 0 |
| Malawi | 243 | 0 | 0 | 0 | 0 |
| Mali | 5 | 6 | 2 | 1 | 36 |
| Mauritania | 10 | 14 | 16 | 0 | 4 |
| Morocco | 51 | 3 | 0 | 0 | 0 |
| Mozambique | 125 | 2 | 1 | 0 | 0 |
| Namibia | 107 | 0 | 0 | 0 | 0 |
| Niger | 3 | 5 | 5 | 0 | 23 |
| Nigeria | 138 | 142 | 168 | 39 | 282 |
| Republic of the Congo | 12 | 0 | 36 | 0 | 0 |
| Rwanda | 30 | 0 | 0 | 0 | 0 |
| Senegal | 0 | 0 | 7 | 7 | 31 |
| Sierra Leone | 5 | 3 | 6 | 0 | 0 |
| Somalia | 37 | 37 | 0 | 0 | 0 |
| South Africa | 51 | 1 | 0 | 0 | 0 |
| South Sudan | 0 | 0 | 6 | 0 | 39 |
| Sudan | 32 | 12 | 0 | 12 | 23 |
| Tanzania | 153 | 16 | 7 | 0 | 0 |
| Togo | 0 | 20 | 9 | 4 | 7 |
| Tunisia | 265 | 1 | 1 | 0 | 0 |
| Uganda | 101 | 33 | 21 | 4 | 4 |
| Western Sahara | 4 | 0 | 0 | 0 | 0 |
| Zambia | 115 | 0 | 0 | 0 | 0 |
| Zimbabwe | 91 | 0 | 0 | 0 | 0 |
| **Total** | **4162** | **637** | **804** | **146** | **601** |

We used a receiver operator curve to understand our model’s performance regarding sensitivity and specificity independent of a single probability cutoff (Fig 5). The area under the curve represents the model’s ability to distinguish between true positives and false positives across all possible classification thresholds. Relative to our model the epidemic risk assigned was higher for districts with epidemics than for those without in 90.8% of ADMN2 districts. In using a probability cutoff value of >0.4 for predicting epidemic experience, the model had a specificity and sensitivity of 80.9% and 84.3%, respectively; these statistics were confirmed in the validation process (Table 4). Here specificity and sensitivity refer to the model’s ability to correctly identify ADMN2 districts that have never/ever been affected by a meningitis epidemic, as in true negative and true positives. As a comparison, Molesworth et al achieved a relative specificity and sensitivity of 67% and 83% within their full model using a cut-off value of >0.4

**Figure 5:** Receiver Operating Characteristic Curve for climatic logistic regression.

**Table 4:**
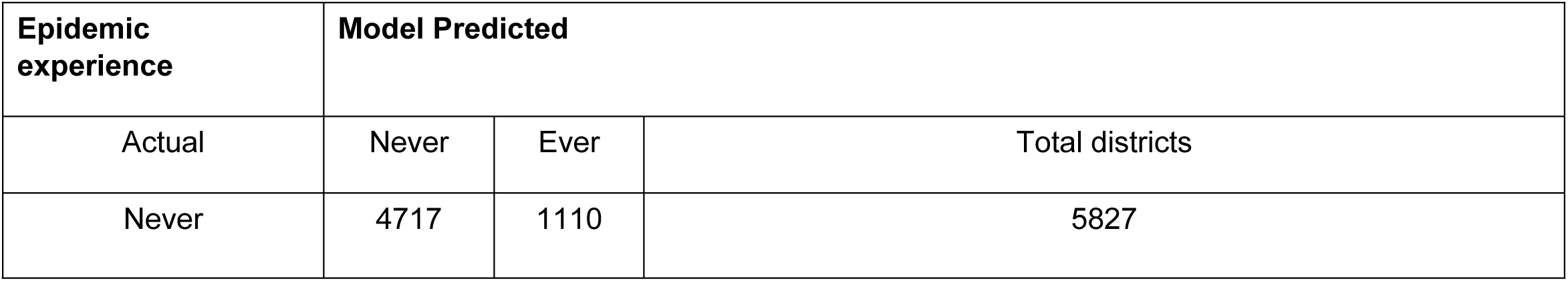
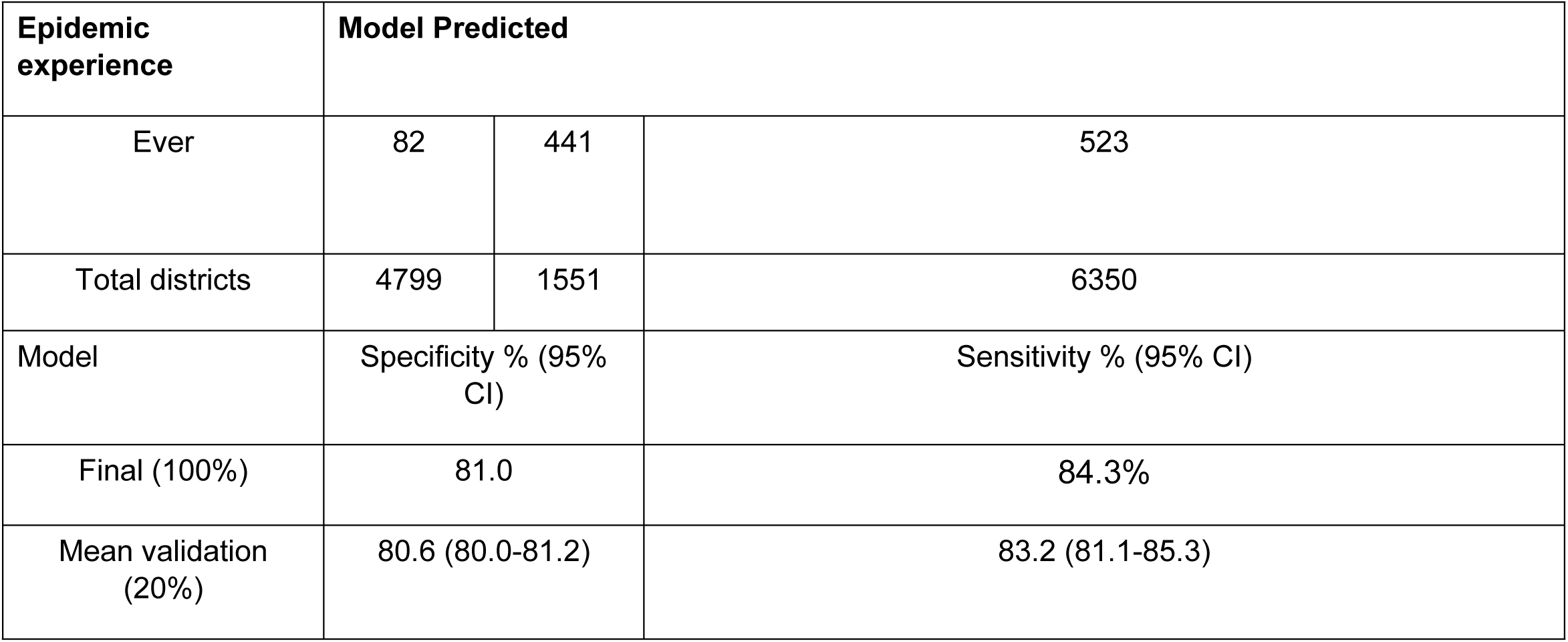
Results of sensitivity and specificity of rainfall, aerosol optical depth, and specific humidity model for both the full dataset and the mean validation using an 80%/20% train and test split.

The results of the logistic regression model without DRC can be found in the Supplementary Information. These results include a full model table (Supplementary Table 1), risk category table (Supplementary Table 2), ROC curve (Supplementary Fig 1) and sensitivity and specificity table (Supplementary Table 3). Overall, removing the DRC improved model fit with a sensitivity of 82.57% and specificity of 91.84%, when using a probability cutoff value of >0.4 for predicting epidemic experience. This is also seen within the ROC curve where epidemic risk assigned was higher for districts with epidemics than for those without in 94.4% of ADMN2 districts.

Within this model, the proportion of districts with a high probability of meningitis outbreaks remained largely comparable to our full model. 9.3% of ADMN2 districts had a very high risk of meningitis outbreaks (probability >0.8) as compared to 9.2% ADMN districts in this risk category in our full model. Burkina Faso, Chad, The Gambia, Mali, Nigeria, Niger, South Sudan and Sudan all had at least 20% of their districts with a very high risk of meningitis outbreaks. Notably, Burkina Faso and Senegal had 91% and 72% respectively of their ADMN2 districts within this risk category and Nigeria had 282 districts classified as very high risk. The key difference within the sensitivity analysis is that a large number of ADMN districts that would have been categorised as moderate risk (probability 0.4-0.59) within the full model that are now low risk (probability 0.2-0.39). Within this model, no country on the periphery of the meningitis belt had any districts with at least a moderate risk of meningitis outbreak (probability 0.4-0.59). Republic of the Congo, Gabon, Liberia, Sierra Leone and Angola which were all at risk of meningitis outbreaks within the full model had no districts with a moderate risk of meningitis outbreaks.

## Discussion

This study examines the spatial patterns of meningitis epidemics through the African continent since 2002 within the context of a changing climate. Our study determined that rainfall, specific humidity, and aerosol optical depth were key predictors in identifying outbreak location. Our model surpasses Molesworth et al’s in environmental raster resolution and has equal if not improved statistical specificity and sensitivity. As in Molesworth et al’s work, districts/regions of Africa with distinct wet and dry seasons were more likely to have meningitis outbreaks. This is highlighted within our rainfall clustering, where Clusters 1 and 2 which had consistently low rainfall from January to March, within the meningitis dry season had the most outbreaks. While there is some variation in aerosol optical depth preceding an outbreak, Cluster 5 which had the second-highest AOD within the typical meningitis dry season had the majority of outbreaks. Both specific humidity and rainfall exhibited seasonal clustering within or around the meningitis belt, emphasizing the climatic influence on meningitis epidemics.

In our full model, we discovered that several countries bordering the meningitis belt to the south were at risk for meningitis outbreaks based on their climate profile. Republic of the Congo, Gabon, Liberia, Sierra Leone and Angola all had at least 15% of their districts with a moderate risk (probability 0.4-0.59) of meningitis outbreaks. Liberia and Angola have both experienced meningitis outbreaks within our study period, suggesting a potential expansion of the region at risk of meningitis [20,35]. This is supported by Savory et al who when evaluating Molesworth’s environmental data with a 100-mile southwards expansion of risk region using data from 2000-2004 were able to improve the pre-existing environmental model sensitivity [20]. Angola has seen an increase in meningitis cases in recent years, most notably within the Huambo province. Between January and May of 2023, the region reported 103 cases, with an estimated lethality rate of 40.8% [36].

Our definition of epidemics was based on countries reporting their data to the WHO-IST in Burkina Faso [26]. This is a well-established system but it is recognized that there is differential reporting between countries. This is a particular issue in the DRC where it is thought that the suspected case definition of bacterial meningitis been poorly applied and where very few suspected cases are tested for laboratoryconfirmation [25].This has had a demonstrable impact on our logistic regression model; when outbreaks from the DRC were taken out of the model as part of a sensitivity analysis specificity increased by 10.94%. Furthermore, when DRC is excluded, there is little evidence for a southerly expansion of the meningitis belt. Within rural ADMN2 districts, suspected meningitis cases are rarely confirmed due to limited laboratory and hospital diagnostic facilities[37]. Improved surveillance of meningitis outbreaks across Africa is vital to validate the potential expansion of high-risk areas and deepen our comprehension of the underlying environmental factors driving these outbreaks. In line with the Defeating Meningitis by 2030 Roadmap, developing reliable and affordable diagnostic tests for bacterial meningitis, could improve case ascertainment within resource-limited settings[38].

Independent of climate change, country-level risk of meningitis outbreaks has also been affected by the introduction of meningitis vaccination programmes. MenAfriVac, offering protection against group A meningococcus, was introduced in mass campaigns across the belt from 2010 onwards, followed by introduction into the essential programme of immunisation (EPI) [39]. Most countries have also introduced pneumococcal conjugate vaccines into EPI over a similar time period. Both vaccines will have influenced the epidemiology of meningitis, with MenAfriVac particularly reducing epidemic risk. However, we excluded vaccination coverage as a risk factor within our model as it will not have an impact on environmental risk factors for meningitis across Africa. Although vaccination will serve to decrease the risk of epidemics in vaccinated areas it will not affect unvaccinated regions and is independent of environmental/climatic variables. Additionally, many outbreaks would have occurred before meningococcal and pneumococcal vaccinations were introduced and the risk of epidemics from other serogroups/serotypes remains

There are some limitations regarding our modelling approach, the most pertinent being our choice to only use environmental predictors. These provide necessary but not sufficient conditions for an outbreak. Several risk factors such as vaccination rates (discussed above), carriage rates of different meningitis-causing bacteria, population-level immunity and physical contact rate have a key role within the dynamics of bacterial meningitis outbreaks across Africa [14,40]. Whilst we used static landcover as a risk factor within our model, further work could consider this as a temporal variable due to the impact of deforestation and desertification on outbreak risk[41,42]. Gridded raster data with long-lat projections used within environmental analysis have unequal cell sizes as longitudinal degrees decrease in length moving outwards from the equator. This means that environmental data in the northern and southern regions of Africa had more granularity than regions at the equator[43]. This was somewhat accounted for within our model through using a cosine of latitude weighting. As in Molesworth et al’s analysis, whilst we received epidemic reports from enhanced disease surveillance countries, other countries outside this region may have unreported meningitis outbreaks. Through combining local data into ADMN2/ADMN1 statistics there is the potential for some discrepancy between regions where patients develop bacterial meningitis and the location of health facilities reporting cases, leading to a loss of data specificity[18]. Finally, the use of k-means clustering and PCA simplifies the data being used, potentially compromising the predictive power of the logistic regression model, if key variations in the data are not captured.

Our model took a 20 year time period as such length is typical in climate analyses. However, we assumed that climatic conditions were static across the reporting period. This may not be the case. Nicholson et al determined that there has been a decrease in precipitation levels preceding the typical dry season across most countries within the northern and western equatorial regions of Africa across the late 20th century. Both areas include countries within the meningitis belt alongside regions identified as having a moderate epidemic risk within our research [44]. Whilst surface temperatures across Africa have been steadily increasing, this is particularly notable in the Sahel where temperatures are rising 1.5 times faster than the global average and is compounded by a shrinking wet season [45]. Ghanian focus group discussion has highlighted participants’ experience of an expanding dry season which now stretches from February to June, with air temperatures rising to 48°C. Notably, there is significant yearly variation within the dry season, potentially accounting for temporal variation in meningitis epidemic burden [46,47]. It has also been suggested that there has been a global increase in the severity and intensity of sand and dust storms including the Harmattan winds over Africa in recent decades, contributing to Sahelian drought persistence and increasing the risk of meningitis outbreaks [10]. Further work will attempt a spatial and temporal analysis to make full use of all available data.

## Conclusion

The environmental risk factors for meningitis epidemics remain largely unchanged across a 40 year period, comparing this analysis to the prior analysis by Molesworth et al. Although our logistic regression model suggests a potential expansion of the meningitis belt, this is dependent on the inclusion of data from DRC, where we know that few cases are laboratory confirmed. Whilst Republic of the Congo, Gabon, Liberia, Sierra Leone and Angola had a moderate risk of meningitis (probability 0.4-0.59) within our full model, no countries surrounding the meningitis belt had a demonstrated risk of meningitis within the sensitivity analysis that excluded DRC. Nevertheless there are clear and measurable associations between epidemic meningitis risk and climate which can be used to inform disease control efforts.

## Data Availability

Epidemiological outbreak data supporting this analyis is available on request. All enviromental data used within this analysis is from publically avaiable sources. R and Stata code used to run this analysis can be found within the following GitHub repository (https://github.com/molly-cliff/Meningitis-belt-location).

## Supporting information captions

S1 Table: Logistic regression results regarding association between meningitis outbreaks and rainfall, aerosol optical depth, and specific humidity

S2 Table: Number of districts within each risk category for all countries across Africa excluding the DRC within the sensitivity analysis.

S1 Figure: Receiver Operating Characteristic Curve for climatic logistic regression sensitivity analysis without the DRC

S3 Table: Results of sensitivity and specificity of rainfall, aerosol optical depth and absolute humidity model for both the full dataset and the mean validation using an 80%/20% train and test split.

---

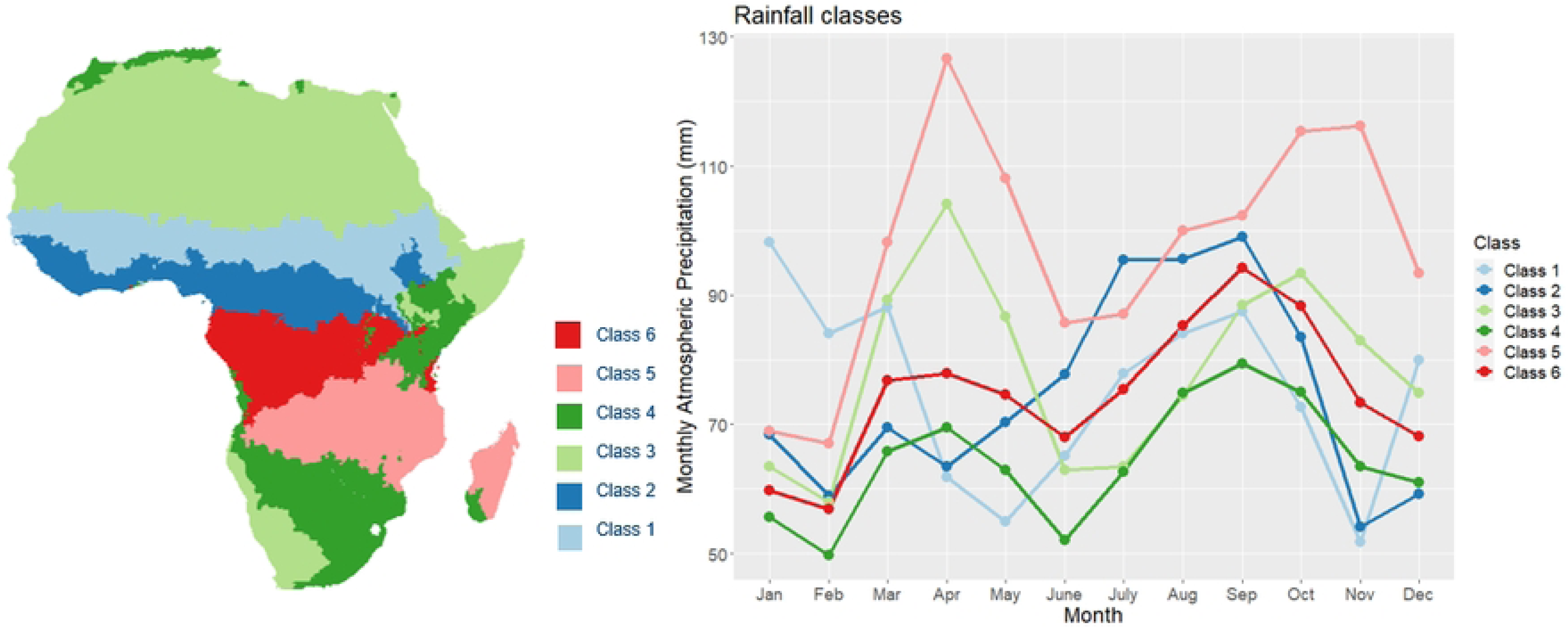

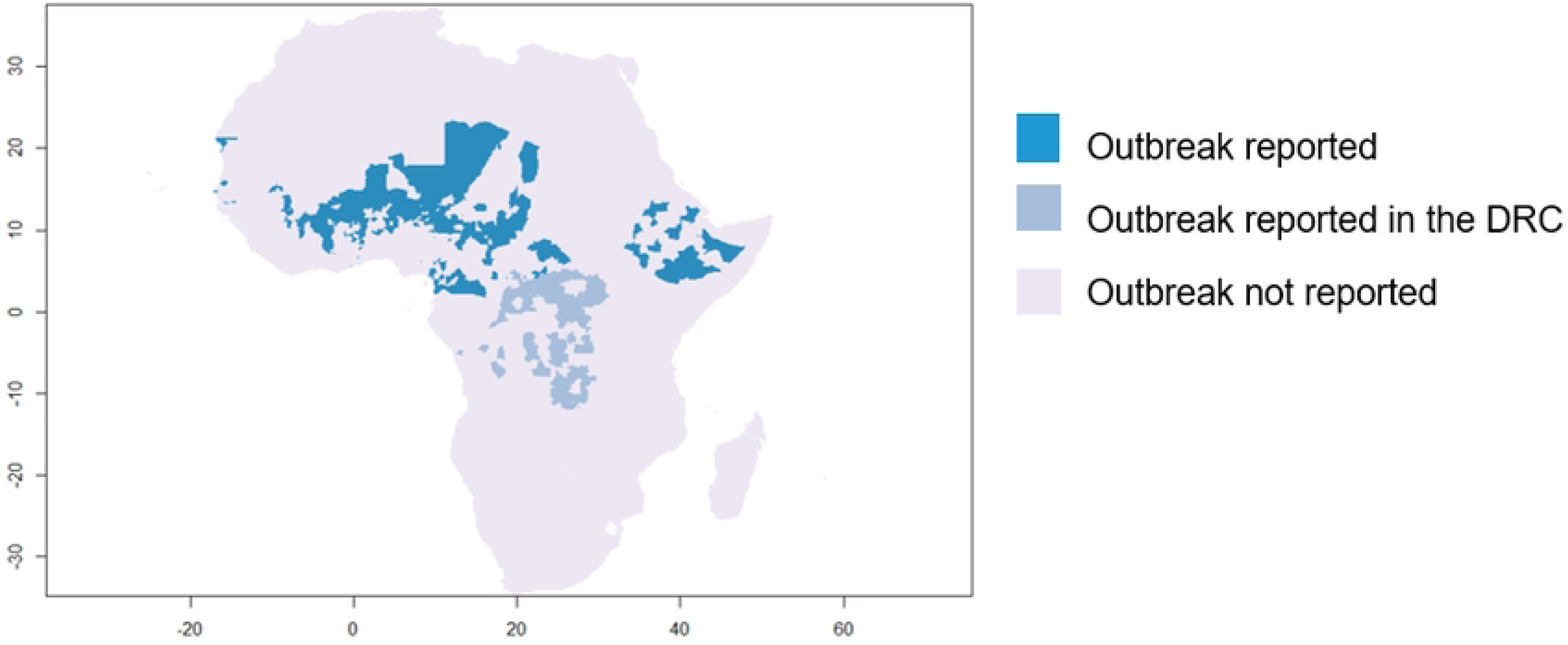

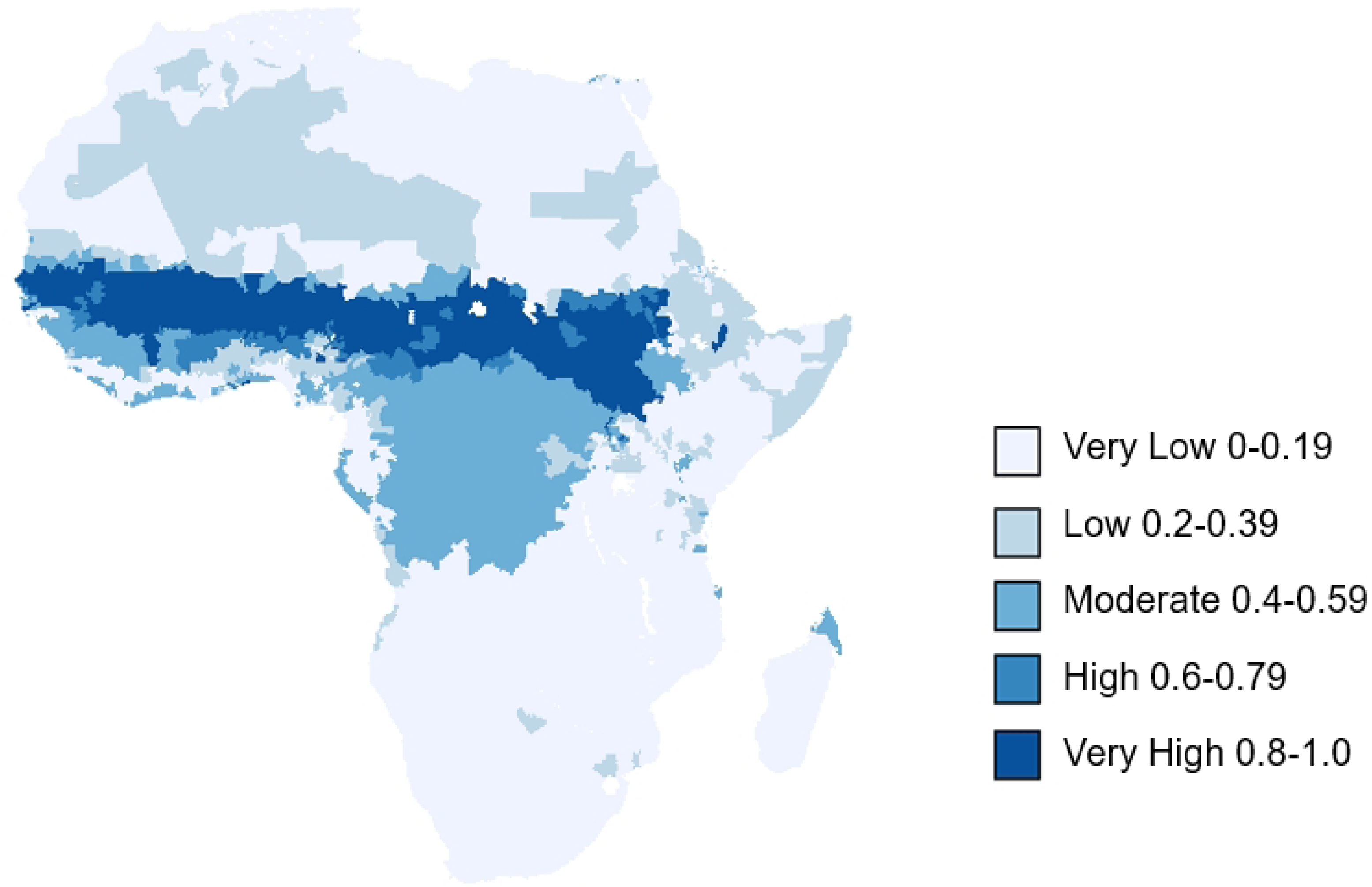

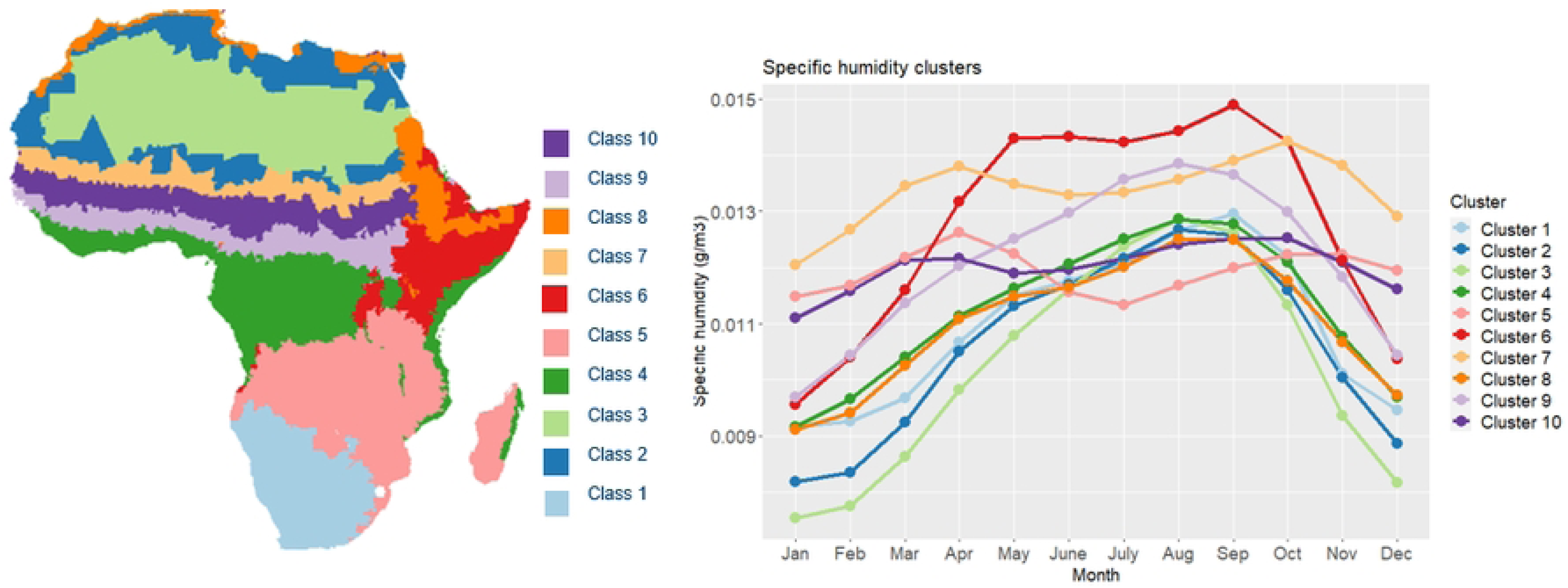

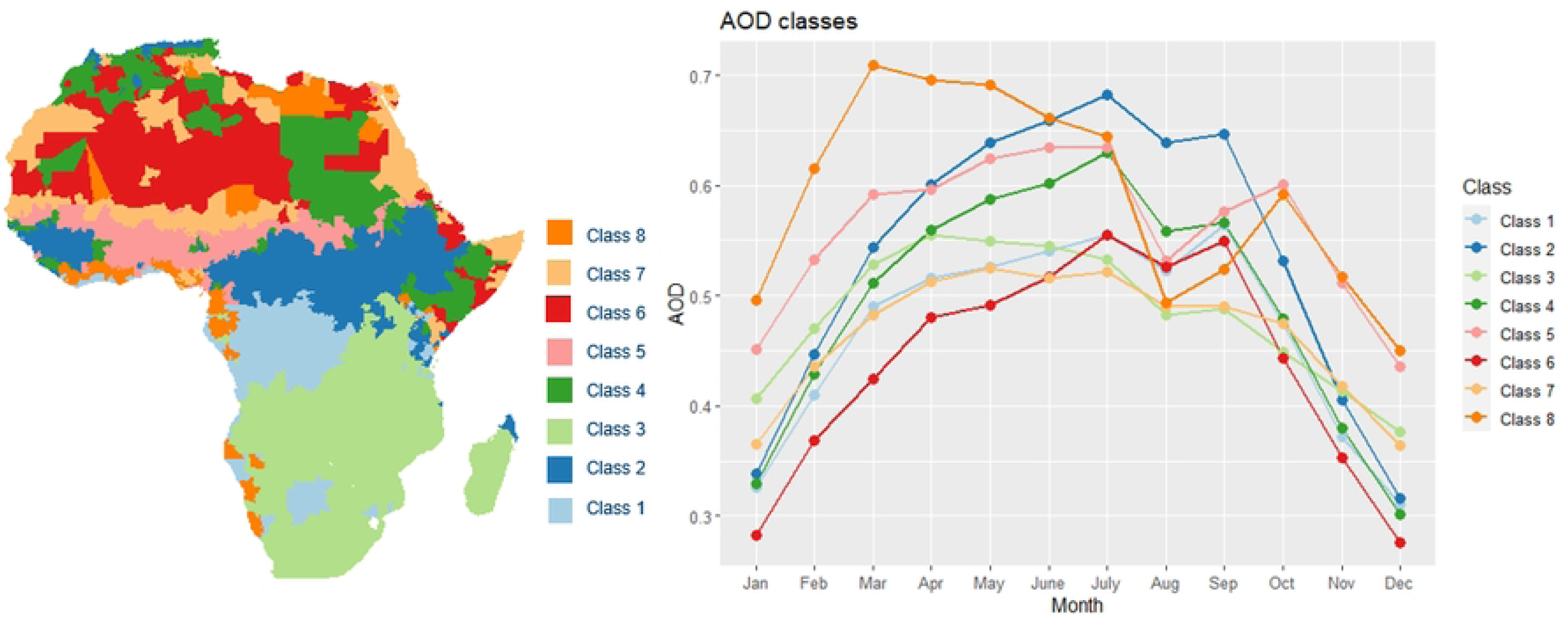

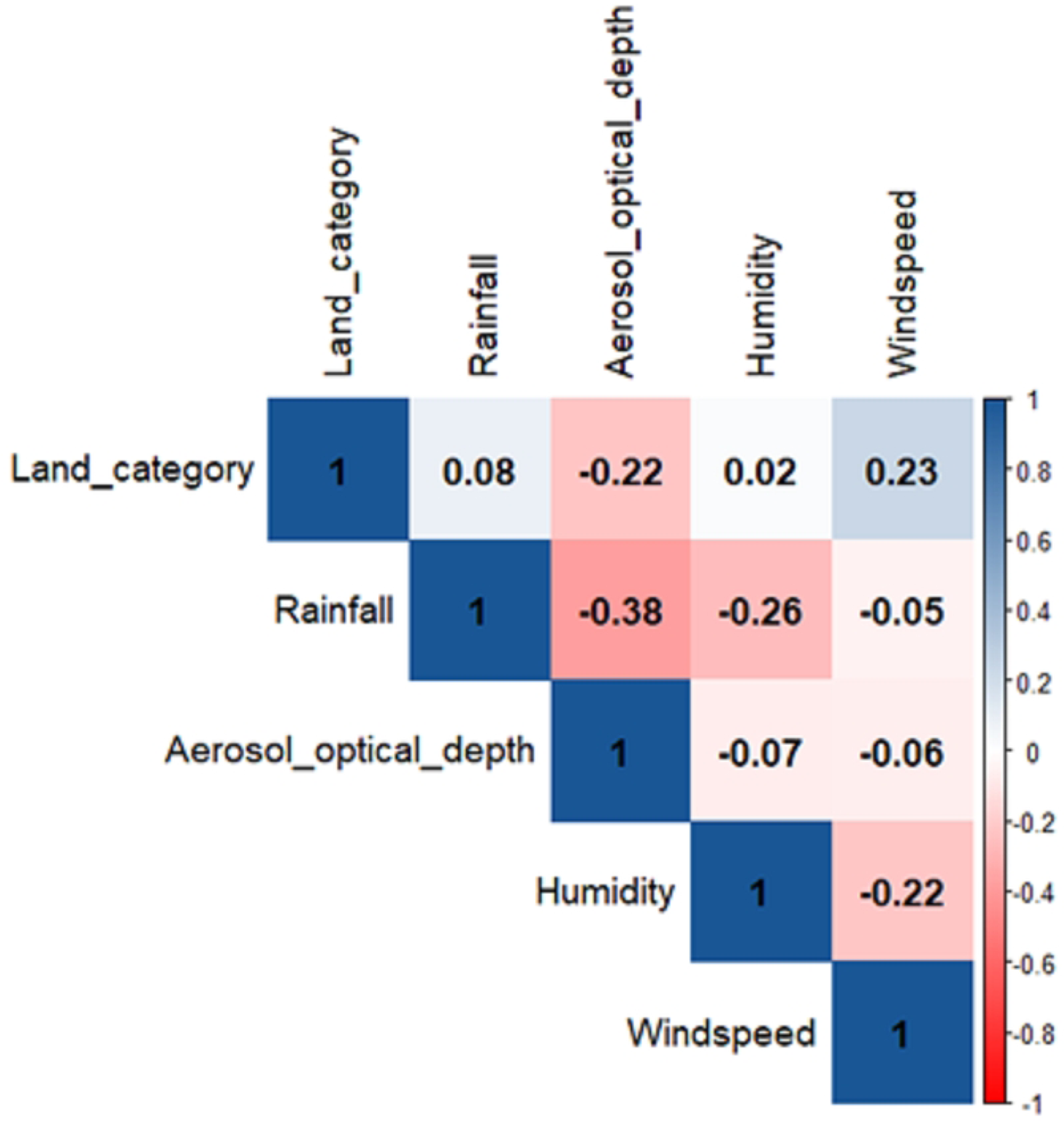

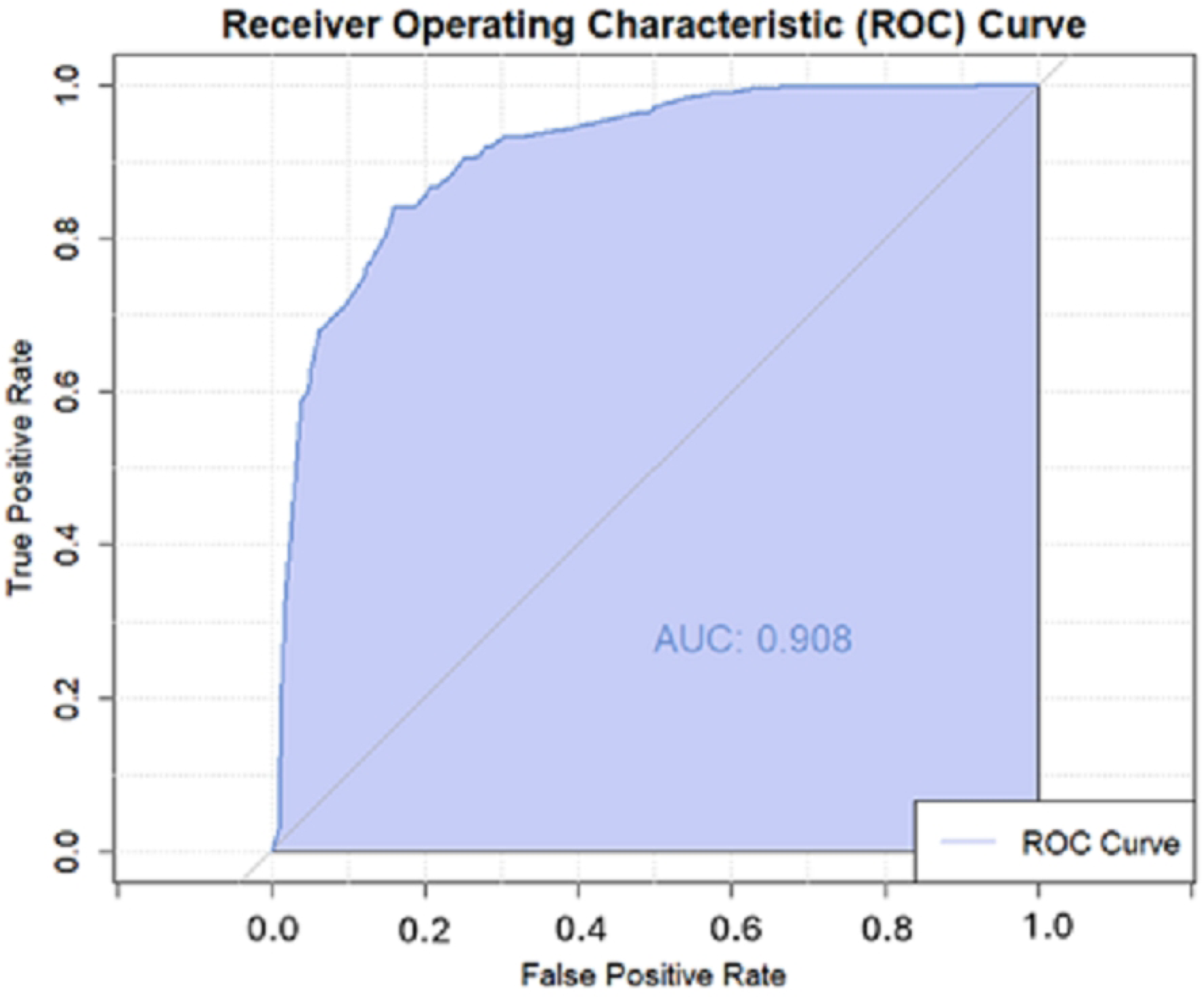

## Notes

### Competing Interest Statement

The authors have declared no competing interest.

### Funding Statement

This work was carried out as part of the Vaccine Impact Modelling Consortium (www.vaccineimpact.org), but the views expressed are those of the authors and not necessarily those of the Consortium or its funders. This work was supported by the Wellcome Trust via the Vaccine Impact Modelling Consortium [Grant Number 226727_Z_22_Z].

### Author Declarations

Ethical approval was not required for this research

